# Bovine Colostrum Derived Antibodies Against SARS-CoV-2 Show Great Potential to Serve as a Prophylactic Agent

**DOI:** 10.1101/2021.06.08.21258069

**Authors:** Kadri Kangro, Mihhail Kurašin, Kiira Gildemann, Eve Sankovski, Eva Žusinaite, Laura Sandra Lello, Raini Pert, Ants Kavak, Väino Poikalainen, Lembit Lepasalu, Marilin Kuusk, Robin Pau, Sander Piiskop, Siimu Rom, Ruth Oltjer, Kairi Tiirik, Karin Kogermann, Mario Plaas, Toomas Tiirats, Birgit Aasmäe, Mihkel Plaas, Dagni Krinka, Ene Talpsep, Meelis Kadaja, Joachim M. Gerhold, Anu Planken, Andres Tover, Andres Merits, Andres Männik, Mart Ustav, Mart Ustav

**Author notes:** **Corresponding author:** Mart Ustav.

## Abstract

Severe acute respiratory syndrome Coronavirus 2 (SARS-CoV-2) until now imposes a serious burden to health systems globally. Despite worldwide vaccination, social distancing and wearing masks, the spread of the virus is still ongoing. One of the mechanisms how neutralizing antibodies (NAbs) block virus entry into cells encompasses interaction inhibition between the cell surface receptor angiotensin-converting enzyme 2 (ACE2) and the spike (S) protein of SARS-CoV-2. SARS-CoV-2 specific NAb development can be induced in the blood of cattle. Pregnant cows produce NAbs upon immunization, and antibodies move into the colostrum just before calving. Here we immunized cows with SARS-CoV-2 S1 receptor binding domain (RBD) protein in proper adjuvant solutions, followed by one boost with SARS-CoV-2 trimeric S protein, and purified immunoglobulins from colostrum. We demonstrate that this preparation indeed blocks interaction between the trimeric S protein and ACE2 in different *in vitro* assays. Moreover, we describe the formulation of purified immunoglobulin preparation into a nasal spray. When administered to human subjects, the formulation persists on the nasal mucosa for at least 4 hours as determined by a clinical study. Therefore, we are presenting a solution that shows great potential to serve as a prophylactic agent against SARS-CoV-2 infection as an additional measure to vaccination and wearing masks. Moreover, our technology allows for a rapid and versatile adaption for preparing prophylactic treatments against other diseases by using the defined characteristics of antibody movement into the colostrum.

**Significance:** SARS-CoV-2 infections continue to be a high-risk factor for mankind. Antibodies with the potential to neutralize the virus and thus its entry into the host cell have been shown to impose a potent measure against the infection. Human derived neutralizing antibodies are therapeutics and thus fall under the legislation of drugs. However, an alternative could be the purification of efficient neutralizing antibodies from other species. Here, we present immunization of pregnant cows with spike protein of SARS-CoV-2 which results in high quantities of colostrum immunoglobulins that can be easily harvested and safely purified within a remarkably short time. The colostrum immunoglobulin preparation has a great potential to serve in formulations that can be used as prophylactic agent against SARS-CoV-2 infection.

## Introduction

Severe acute respiratory syndrome Coronavirus 2 (SARS-CoV-2) emerged in the Chinese province of Hubei in December 2019 and spread worldwide within few months leading to declaration of a pandemic in March 2020. The disease caused by SARS-CoV-2 was named COVID-19 by the World Health Organization (1). Signs and symptoms of COVID-19 may range from very mild to severe and appear 2 to 14 days after exposure. Symptoms include runny nose, sore throat, dry cough, muscle/joint pain, loss of taste/smell, shortness of breath, fever, chills/shaking, diarrhea, nausea/vomiting, fatigue and/or headache (2). The impact of the SARS-CoV-2 on the world-wide healthcare systems and economy has been devastating. Even with multiple vaccines on the market by now, the treatment and prevention of the SARS-CoV-2 infection are still to be developed. Moreover, the virus is continuously adapting, and new viral variants might escape recognition by vaccine-induced immunity (3). The most notable variants of concern (VOC) have emerged in United Kingdom (Alpha, B.1.1.7), South Africa (Beta, B.1.351), Brazil (Gamma, P.1) and more recently in India (Kappa, B.1.617.1 and Delta, B.1.617.2) (4–6).

SARS-CoV-2 belongs to the family *Coronaviridae* that are positive-sense single-stranded RNA (+ssRNA) viruses, and is a member of the subgenus *Sarbecovirus* (*Betacoronavirus* lineage B) (1). Like most of other coronaviruses, SARS-CoV-2 virion contains four structural proteins: S (spike), E (envelope), M (membrane), and N (nucleocapsid) proteins. N protein binds the RNA genome, S, E, and M proteins are localized in viral envelope (7). The trimeric S protein comprises monomers that consist of S1 and S2 subunits, and it facilitates the entry of the virus into the host cell (8). More specifically, the receptor binding domain (RBD) in the S1 subunit is responsible for the recognition and binding to the angiotensin-converting enzyme 2 (ACE2) (9) on a host cell, which is followed by proteolytical activation by host proteases (10). Thereafter, the S2 subunit mediates the fusion between the virion envelope and the membrane of the host cell (11). ACE2 is abundantly localized in the epithelia of the lung and small intestine which provides viral entry into human cells (12, 13).

Neutralizing antibodies (NAb) have been found to block the entry of a pathogen into the cell and thus prevent infection (14, 15). Moreover, SARS-CoV-1 anti-S antibodies have been shown to play a major role in blocking the virus entry in a hamster model, while high titers of anti-N antibodies did not provide any protective immunity (16). Thus, since the initial encounter between the virus and the host is mediated by the RBD region, the majority of the NAbs are directed against RBD (17), although, in some cases the NAbs might target other epitopes on the trimeric S outside the RBD region as well (18). Therefore, finding efficient NAbs that could block the entry of SARS-CoV-2 provides a promising approach for developing prophylactic and/or therapeutic means to fight against the pandemic. Passive immunization (19) with anti-SARS-CoV-2 NAb could be especially valuable for certain populations that are suffering the most: the elderly, the immunocompromised, patients in nursing homes and long-term care facilities, and chronically ill patients.

The passing of protective immunity through colostrum in mammals is a naturally evolved process that provides protection against exogenous pathogens to the newborn (20–22). Ungulates cannot transfer immunoglobulins across the placenta, and the intestine of a newborn is permeable to proteins only up to 24 hours after birth (20). Consequently, the colostrum has to contain elevated levels of immunoglobulins that form up to 70-80% of the total protein (23). The maternal serum immunoglobulins decrease rapidly before parturition, and are transported into the colostrum (24) with the main immunoglobulin class being IgG (up to 90%), followed by IgM and IgA (21). The IgG levels in bovine colostrum are found to reach up to 20-200 mg/mL (21, 25, 26).

The use of bovine colostrum as a food supplement to humans (26–28) has been demonstrated to elicit beneficial effects against intestinal pathogens (21). Therefore, the development of hyperimmune bovine colostrum can be an excellent source of specific antiviral antibodies. It has been found that the intranasal administration of colostrum antibodies from cows who were vaccinated with influenza vaccine, could protect mice from development of infection when a sublethal dose was administered (29). Similar transfer of hyperimmune colostrum derived protection has also been demonstrated against *Clostridium difficile* infection (30). Moreover, the intranasal administration of SARS-CoV-2 human NAb was shown to prevent infection in mice (31). Therefore, the combination of colostrum derived antibodies from cows immunized with SARS-CoV-2 spike protein in a formulation of an intranasal preparation might provide a powerful, affordable and flexible tool to provide protection in the upper respiratory tract, the portal of entry of SARS-CoV-2 infection. Here we demonstrate that such a colostrum polyclonal antibody preparation can provide efficient block against SARS-CoV-2 virus infection, including several known escape variants (32). We also describe a nasal spray formulation containing the colostrum antibody preparation and its bioavailability in the human nasal cavity.

## Results

### Immunoglobulin preparation from colostrum

Of 8 immunized cows, only 7 cows reached a successful calving, and thus provided colostrum. Colostrum was chymosin treated, and lipids were removed to obtain whey. Since the two different immunization schemes resulted in a rather equal outcome, all the whey fractions containing NAbs were pooled. Whey was further pH treated, underwent different filtration and fractional precipitation steps, and was finally formulated into a buffer solution (Fig. 1).

**Figure 1.**
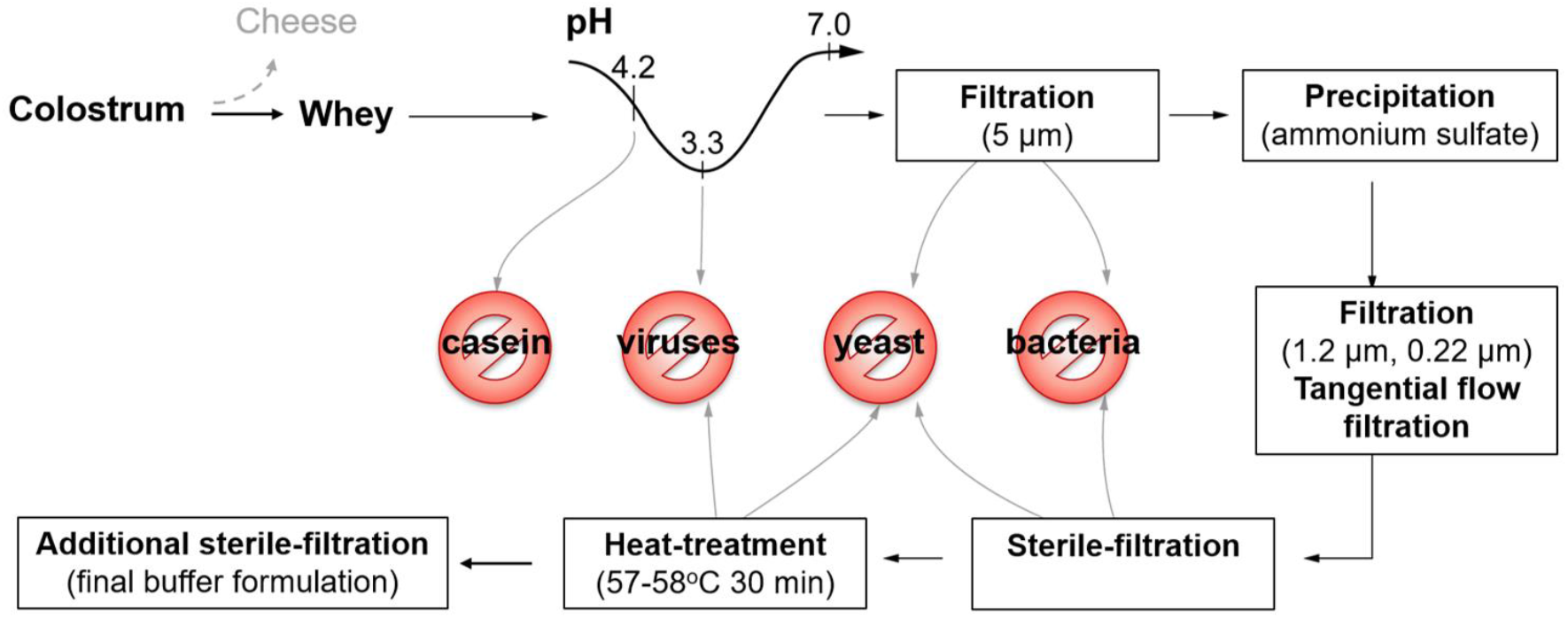
Schematic overview of the bovine immunoglobulin preparation from colostrum.

### Blocking of trimeric S protein and ACE2 interaction by colostrum derived neutralizing anti-S antibodies

To characterize blocking of the interaction between trimeric S and ACE2 by the colostrum immunoglobulin preparation of different SARS-CoV-2 variants, we determined the interaction blocking efficacy of the antibody preparation using a competitive enzyme-linked immunosorbent assay (ELISA). The colostrum immunoglobulin preparation was added in a serial dilution to the pre-coated trimeric S proteins of the wild type (wt), Alpha, Beta, Gamma, Delta or Kappa VOCs, followed by incubation with conjugated ACE2. Since the antibodies and enzyme conjugate are competing for the same binding sites (Fig. 2A; left panel), the binding rate of the enzyme conjugate revealed whether the colostrum immunoglobulin preparation was able to block the interaction between ACE2 and trimeric S. Relative OD values ≥0.75 were determined to show no ACE2 blocking capacity by the antibodies, since the conjugated ACE2 could still bind to the RBD site. Relative OD values <0.75 indicate ACE2 blocking capacity of the analyzed antibody, as the antibodies could bind to the coated S protein and thereby hindering the further binding of ACE2 to the RBD. The half-maximal inhibitory concentration (IC_50_) was calculated for the binding to each trimeric S protein variant. We found that colostrum immunoglobulin preparation blocks the ACE2 and wt, Alpha, Beta, Gamma, Delta or Kappa trimeric S interaction with an IC_50_ of 109.7 µg/mL, 250.2 µg/mL, 105.5 µg/mL, 300.9 µg/mL, 97.98 µg/mL and 136.8 µg/mL, respectively (Fig. 2A; right panel). Thus, the colostrum immunoglobulin preparation should be able to block the viral entry into human cells.

**Figure 2.**
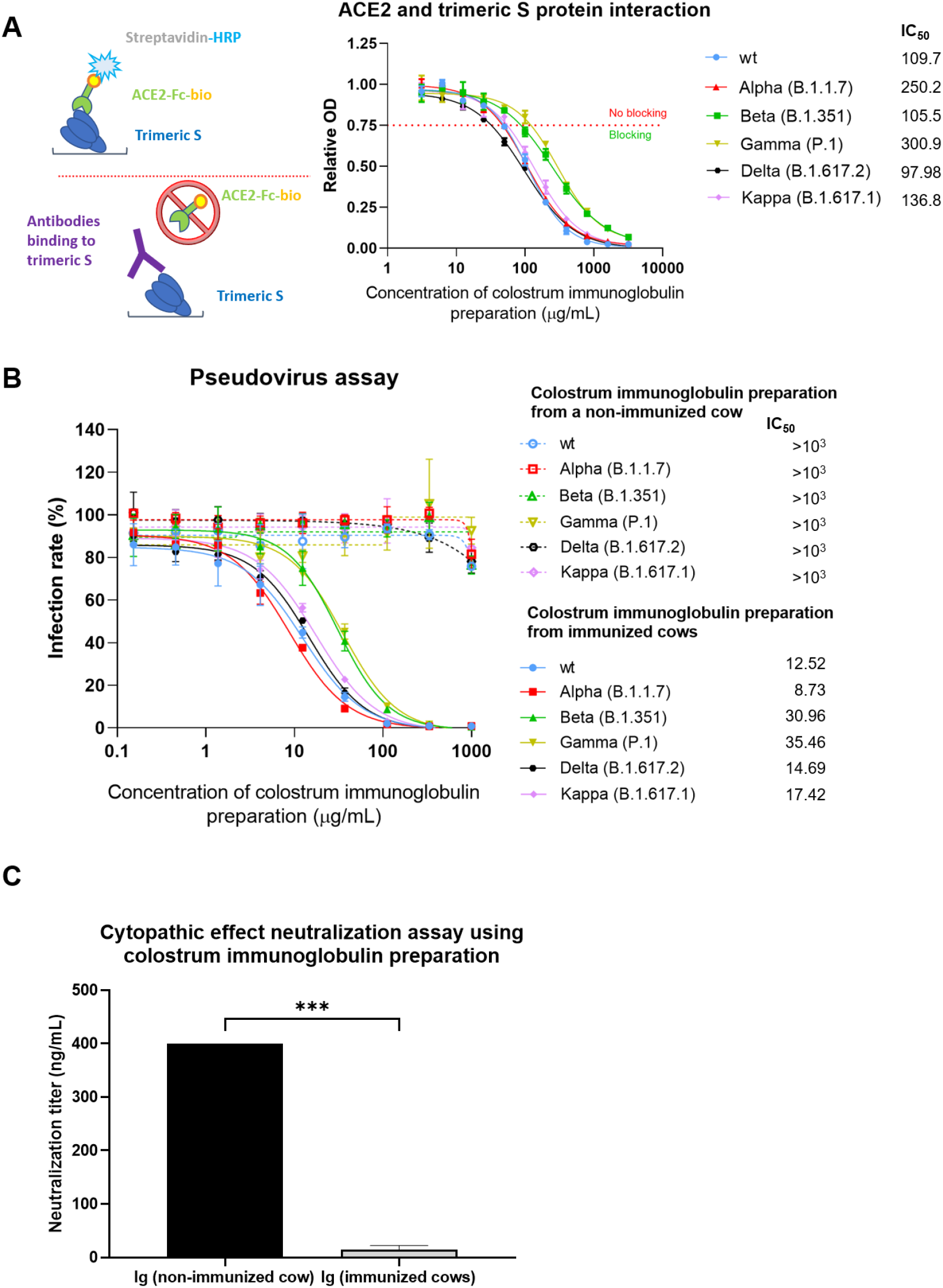
Functional characteristics of SARS-CoV-2 NAbs from colostrum. (A) Left panel: NAb ELISA assay set-up and principle. Colostrum immunoglobulin preparation was added in serial dilution to the trimeric S protein coated plates and pre-incubated in order to provide some time for the antibodies to bind. Next, the conjugated ACE2 which is competing with the antibodies for the same binding sites, was added. Coloring development becomes more intense in samples containing fewer neutralizing antibodies, and a sample without any antibodies has the most intense color, since maximal binding of conjugated ACE2 could occur. Right panel: Binding of the colostrum immunoglobulin to the different trimeric S protein variants in SARS-CoV-2. Relative optical density (OD) values are represented as mean ± SD (n=3). The IC_50_ concentrations for colostrum immunoglobulin preparation were determined using 4-parameter nonlinear regression. (B) Pseudovirus neutralization assay. Measurement of the inhibition of trimeric S protein dependent entry of different pseudoviruses that were pseudotyped with either wt, Alpha, Beta, Gamma, Delta or Kappa VOCs of the SARS-CoV-2 S protein into ACE2 expressing HEK293 cells in the presence of serial dilutions of colostrum immunoglobulin. Rate of cell entry was measured via firefly luciferase activity, and expressed as relative infection rate (%) compared to luciferase activity in untreated control. (C) The neutralizing activity of the colostrum immunoglobulin on SARS-CoV-2 infection analyzed by cytopathic effect reduction. Colostrum derived immunoglobulins (Ig) from a non-immunized cow and from SARS-Co-V-2 S-protein immunized cows were added to SARS-CoV-2 virus isolate at different concentrations and then transferred to Vero E6 cells which were grown for 4-5 days. The neutralization activity was defined by endpoint method *i*.*e*. determination of colostrum immunoglobulin preparation concentrations blocking infection of the Vero E6 cells.

### Pseudovirus neutralization assay

To further characterize the virus neutralizing potency of the colostrum immunoglobulin preparation, a pseudoviral neutralization assay was performed in the presence of serial dilutions of immunoglobulin preparations. ACE2 expressing human embryonic kidney (HEK)293 cells were infected with a HIV-based pseudovirus delivering a luciferase marker gene to the infected cells. These viral particles were pseudotyped with either wt, Alpha, Beta, Gamma, Delta or Kappa VOC of the SARS-CoV-2 S protein. We found that the colostrum immunoglobulins from the non-immunized cow showed no effective inhibition even at highest concentration (IC_50_ >10^3^ µg/mL). In contrast, immunoglobulins from immunized cows blocked the entry of pseudoviruses carrying the S protein of either wt, Alpha, Beta, Gamma, Delta or Kappa into the cells with an IC_50_ of 12.52 µg/mL, 8.73 µg/mL, 30.96 µg/mL, 35.46 µg/mL, 14.69 µg/mL and 17.42 µg/mL, respectively (Fig. 2B).

### SARS-CoV-2 induced cytopathic effect neutralization assay

NAbs can have additional activities (*e*.*g*., act via epitopes outside of receptor binding domain of the S protein) useful for the inhibition of the viral infection. Thus, we also tested the inhibitory effect of the colostrum immunoglobulin preparation using authentic virus isolated from an individual who recovered from COVID-19, and Vero E6 cells which are highly susceptible for SARS-CoV-2 infection resulting in prominent cytopathic effects (morphology change, detachment from the substrate). An immunoglobulin preparation from a non-immunized cow failed to protect cells at concentrations lower than 400 ng/mL (Fig. 2C). The protective concentration of the colostrum immunoglobulin preparation from immunized cows was observed on average at 14.8 ng/mL (Fig. 2C), which is more than 25-fold lower (p=0.0005) compared to the protective concentration of the control colostrum.

### Characteristics of immunoglobulin preparation in the nasal spray formulation

For practical use as preventive means, the antiviral formulation preferably has to be maintained on the site of action for several hours, and the nasal spray dispenser can be used for effective administering solutions to the upper respiratory tract. To establish the recommended dosage regimen for use and further efficacy studies, we determined how long the immunoglobulin preparation formulation remains on the nasal mucosa. Sixteen healthy volunteers were recruited in a clinical trial. The immunoglobulin preparation as formulated into a nasal spray was administered at two different concentrations by spraying twice into each nostril. Samples were taken one hour and four hours after the administration to measure the residual bovine immunoglobulin G on the nasal cavities. In the baseline samples collected prior to administration, no bovine IgG was detected. In the subgroup of the lower dose (0.1 mg/mL), one hour and four hours after the administration, the average bovine IgG concentrations were 1.54 µg/mL and 1.16 µg/mL, respectively (Fig. 3). As expected, in the subgroup of the higher administered dose, the bovine IgG were detected at concentrations of 5.65 µg/mL and 2.36 µg/mL, respectively (Fig. 3). Thus, even at the later time point, the preparation was clearly detectable. No adverse effects were reported by the participants in either of the study groups. Therefore, we concluded, that although, the bovine antibodies on the nasal mucosa were detectable when applying different doses of colostrum immunoglobulin preparation, a higher concentration is more preferred as it allows more convenient dosage frequency after every 3-4 hours to retain a relevant concentration of antibodies on nasal mucosal surfaces.

**Figure 3.**
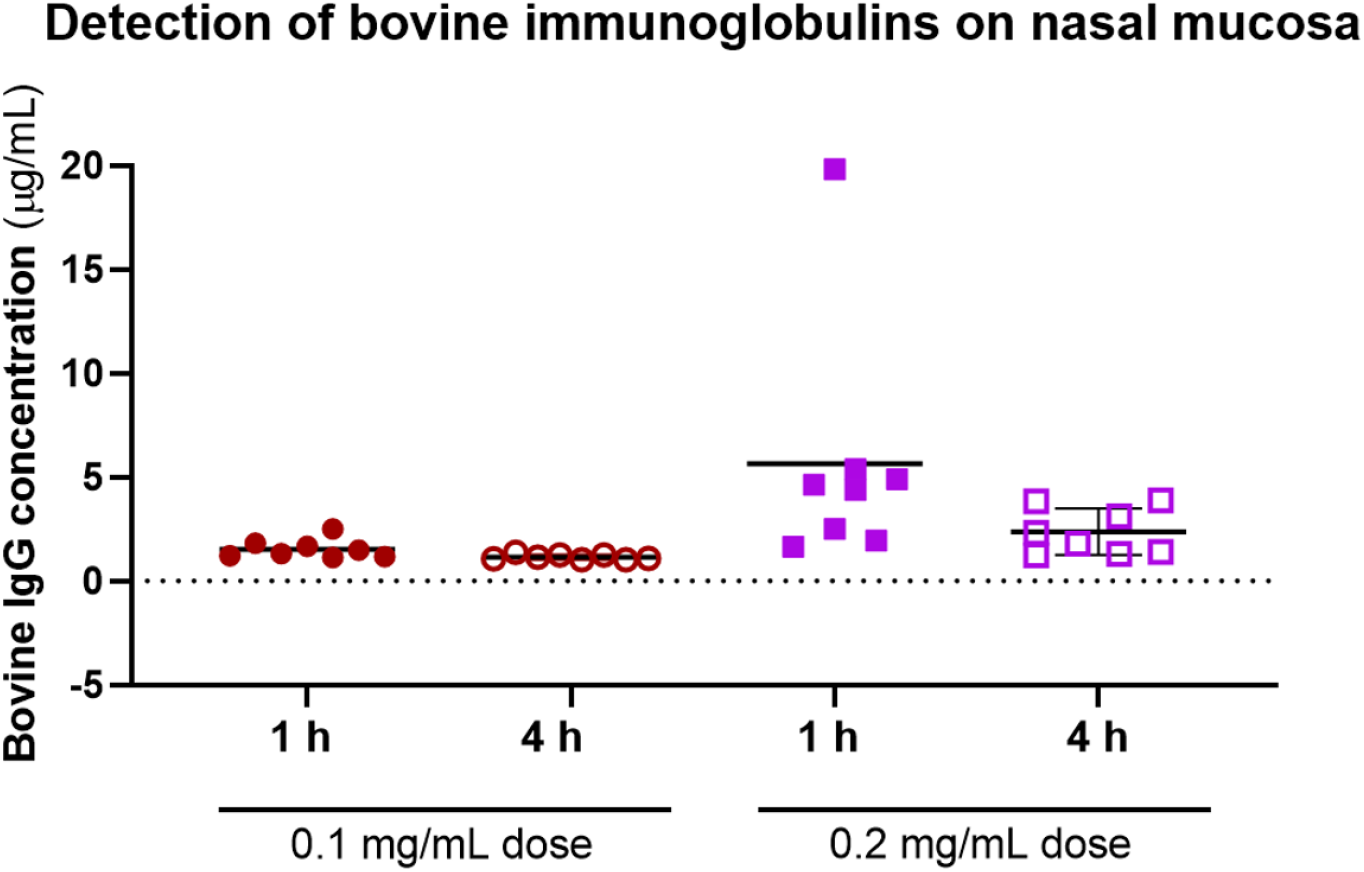
Bioavailability of the final immunoglobulin formulation which was administered intranasally to healthy volunteers at two different concentrations (0.1 mg/mL and 0.2 mg/mL) and the residual solution 1 h and 4 h post-administration was measured with a cow IgG ELISA.

## Discussion

As vaccines are providing protection from the development of lethal COVID-19 (33), the need for additional preventive measures to complement the vaccines to reduce the infection and spread of SARS-CoV-2 are in dire need. The development and emergency use authorization of monoclonal antibodies for an early infection stage therapeutic and preventive approach has proven to be effective (34). Issues related to cumbersome administering and immune evasion of the emerging VOC have reduced the widespread implementation of these therapies. Further studies have indicated that COVID-19 illness has distinctive stages, with indicative clinical findings. The primary infection occurs in the upper respiratory tract epithelium, followed by the second stage of pulmonary disease with viral multiplication and inflammation in the lungs (35). By providing an environment in the upper respiratory tract, mucosal surfaces that contain virus neutralizing antibodies can be an attractive strategy for providing protection against the early-stage infection. Here we show that a novel nasal spray containing bovine colostrum derived antibodies against SARS-CoV-2 shows great potential to serve as a prophylactic agent. This adds to the range of measures that are taken or can be taken to fight the spread of SARS-CoV-2.

We demonstrated that immunization of Estonian Holstein breed cows leads to the production of NAbs in their blood serum and that these antibodies efficiently move into the colostrum from where these antibodies can be easily purified (Fig. 1). Using a competitive ELISA, which measures the ability of potentially neutralizing antibodies to block interaction between the RBD on the trimeric S protein and ACE2, we showed that this interaction is not only inhibited by the colostrum derived immunoglobulin preparation, but that this polyclonal antibody cocktail is also efficient towards different VOCs (Fig. 2). This observation was further confirmed in a pseudovirus assay, indicating that the polyclonal antibody preparation from colostrum reveals a broad enough specificity against currently widely spread SARS-CoV-2 VOCs. These results furthermore underscore the great potential of the bovine colostrum immunoglobulin preparation as a promising solution to inhibit virus spread in addition to vaccination. Moreover, the colostrum immunoglobulin preparation remains on the human nasal mucosa for at least 4 hours, as the formulation contains viscosity increasing excipients (see the M&M section for details) to prolong the residence and action time on the nasal mucosa, and hence assures the successful local delivery of antibody preparation. Preliminary data of anti-S antibody concentrations on the nasal mucosa in patients fully vaccinated with mRNA vaccines is in average 2.5 µg/mL (36). From the colostrum derived immunoglobulin formulation as presented here, if delivered twice per nostril at 0.2 mg/mL, a minimal required concentration is retained at a comparable concentration range on mucosal surfaces after 4 hours as detected for fully vaccinated individuals (36). In addition, the measured residual bovine immunoglobins persist on the nasal mucosa in a much higher concentration than the neutralizing capacity of the formulation as measured in our SARS-CoV-2 induced cytopathic effect neutralization assay.

Together we conclude that the colostrum derived immunoglobulin preparation formulation provides a very efficient neutralization effect *in vitro*. Our results may bare high significance since it is being reported that polyclonal antibodies from convalescent patients, vaccinated individuals, and different immunized animals may show reduced neutralizing potency to different VOCs of SARS-CoV-2 (15), but a definite neutralization effect has never been shown. Moreover, this technology of colostrum derived immunoglobulins can be employed for producing (neutralizing) antibodies against various other existing or arising viral diseases and variants thereof.

## Material and Methods

### Immunization of pregnant cows

All animal experimental protocols were approved by the Estonian Project Authorization Committee for Animal Experiments on November 5^th^ 2020 (approval number 177), and all experiments were performed in accordance with the European Communities Directive of September 2010 (2010/63/EU) and the study was carried out in compliance with the ARRIVE guidelines. Eight cows (Estonian Holstein breed) were intramuscularly immunized twice with SARS CoV-2 S RBD protein (0.1 mg of antigen per injection) followed by 1 boost with SARS-CoV-2 trimeric S protein (0.5 mg per injection) in proper adjuvant solutions. For adjuvant, a mixture of Quil-A (0.5 mg/mL, Invivogen) and Imject Alum (20 mg/mL, Thermo) was used for immunizing 3 cows, and Quil-A (0.5 mg/mL, Invivogen) alone was used for immunizing 5 cows. The first immunization was performed 61-65 days (for 4 cows) and 56, 72 and 79 days, respectively for 3 cows, before the expected calving date. The second immunization was performed three weeks after the first injection, and the boost was performed two weeks after the second injection.

### Extraction of antibodies from whey

Twice-daily collected colostrum of each cow was collected separately, and defatted casein-depleted whey was frozen at -20°C and held until purification steps. Different fractions of whey were first analyzed for the presence of NAbs using an ELISA (Icosagen, K5-002-096), and the fractions containing NAbs were pooled for purification. To remove casein remains the whey was melted at +4°C, then pH was adjusted to 4.2-4.5 using 1 M HCl and the whey was left at room temperature for 1 hour with continuous mixing using a magnet stirrer. Thereafter pH was adjusted to 3.3 with 1 M HCl, and the whey was left for 1 hour at room temperature to inactivate possible viral contaminants. The acid pH-treated colostrum whey was neutralized to pH 6.7-7.0 using 1.5 M Tris-HCl pH 8.8 and filtrated through a 5 µm filter. The filtrated solution was mixed with ammonium sulfate to a final concentration of 2 M and incubated at +4°C overnight. The protein precipitate was separated from the supernatant by centrifugation at 7000x g for 15 min at 4°C. The precipitated proteins were dissolved in 1x DPBS (Dulbecco’s phosphate-buffered saline) solution and concentrated to 100 mg/mL. Concentration was determined by measuring the UV absorbance at a wavelength of 280 nm (A^0.1%^ 280 nm = 1.37). The concentrated immunoglobulin-enriched solution was filtered through 1.2 µm a prefilter and a 0.22 µm filter, and dialyzed against 1x DPBS by means of tangential flow filtration (TFF). TFF was carried out on Äkta Flux 6 using Sartocon cassettes (50 kDa cutoff). Dialyzed proteins were sterilized by filtration (0.22 µm filter) followed by pasteurization at 57-58°C for 30 min (37).

### SARS-CoV-2 neutralizing antibody ELISA

The wells of Nunc MaxiSorp flat-bottom 96-well plates were coated with purified SARS-CoV-2 trimeric S protein of different VOCs at 2.5 µg/mL in PBS overnight at 4°C, and blocked with a PBS solution containing 1% bovine plasma albumin (BPLA) for 1 hour at room temperature (RT). The samples were diluted in assay buffer (0.5% BPLA in PBS) and added in a 1-in-2 dilution in triplicates to the blocked plate with a starting concentration of 3.2 mg/mL in a final volume of 50 µL, and pre-incubated for 20 min at RT. On each 96-well plate an assay buffer containing no antibodies was added in three repeats (negative control). Next, an enzyme complex (50 µL) containing biotinylated ACE2-hFc and Pierce™ High Sensitivity Streptavidin HRP (horse radish peroxidase)-labelled at a concentration of 0.5 µg/mL, was added to the pre-incubated plates, and incubated for 30 min at RT. Colorimetric development was performed by using 3,3’,5,5’-tetramethylbenzidine VII substrate. The reaction was stopped using 0.5 M H_2_SO_4_, and absorbance was measured at 450 nm. The OD values of the measured samples were divided by the mean value of the three repeats of negative control to obtain relative OD values.

### Pseudovirus neutralization assay

HEK293 cells were co-transfected via electroporation with ACE2 encoding DNA fragment and Hygromycin encoding DNA, and grown in DMEM in the presence of 200 µg/mL Hygromycin for 2 weeks for selection. ACE2 expressing HEK293 cells were seeded into 6-well plates at 5×10^6^ cells per well. Next day, cells were transfected with a vector, containing wt, Alpha, Beta Gamma, Delta or Kappa VOC of the SARS-CoV-2 S protein. Transfection was carried out with Lipofectamine 3000 according to the manufacturer’s protocol (Thermo). At 72 h post-transfection pseudovirus containing medium was collected, filtered through a 0.45 µm filter, and used immediately to infect ACE2-expressing HEK293 cells. For the pseudovirus neutralization test, 10^5^ ACE2-expressing HEK293 cells were incubated using the pseudovirus-containing medium in the presence of appropriate colostrum immunoglobulin preparation dilution series (from concentration 1 mg/mL to 0.15 µg/mL). On the following day, the culture medium was removed and replaced with DMEM. After incubation for 72 h, the Renilla luciferase activity was measured using a GloMax reader (Promega). Experiments with colostrum immunoglobulin preparation from immunized and non-immunized cows were performed in three and two biological repeats, respectively.

### SARS-CoV-2 induced cytopathic effect neutralization assay

A 3-fold serial dilution of the colostrum immunoglobulin preparation (from concentration 200 ng/mL to 3.3 pg/mL) from immunized cows and from a non-immunized cow was made using VGM (Virus Growth Medium, DMEM supplemented with 0.2% BSA). Medium without the preparation was used as a positive control for infection. The dilutions were made in a 96-well plate in duplicate. Thereafter, 100 PFU of the SARS-CoV-2 (Estonian isolate 3542, Wuhan strain) was added per well and sample was incubated for 1 hour at 37°C. Next, 4x 10^4^ Vero E6 cells were added to each well and the plates were incubated for 4 days at 37°C in humid environment before evaluation of the cytopathic effect caused by viral infection. The same number of the cells were grown without the virus as negative control. After incubation, the cytopathic effect was evaluated microscopically.

### Formulation of bovine immunoglobulin preparation for nasal spray

The bovine colostrum immunoglobulin preparation in a nasal spray formulation contains 0.1% sodium benzoate, 0.07% citric acid, 5% polyethylene glycol 400, 1.5% glycerol and 1% polyvinylpyrrolidone K30 in 1x DPBS. All excipient concentrations were selected based on the formulation studies using various multicomponent formulation compositions. The nasal spray formulations were tested for pH (digital pH meter), viscosity (Brookfield LVDVNX CP Rheometer), particle size and polydispersity index (Malvern Zetasizer), and sterility (Eur.Pharm.10th; 2.6.1). The selection of viscosity enhancers and their concentration was made based on the viscosity and droplet size measurements. For bioavailability study, the colostrum immunoglobulin preparation was prepared at two different concentrations 0.1 mg/mL and 0.2 mg/mL in the nasal spray formulation.

### Bioavailability of colostrum immunoglobulin preparation

To test the nasal biological availability of the bovine colostrum immunoglobulin preparation a clinical study on 16 healthy volunteers was conducted in accordance with the Declaration of Helsinki, approval was granted by the Tartu University Ethics Committee on March 17^th^ 2021 (approval number 336/T-1) and the trial was registered at ClinicalTrials.gov (ClinicalTrials.gov Identifier: NCT04916574). Written informed consent was obtained from each healthy volunteer. The study group was divided into two subgroups where the individuals were intranasally administered (double spraying; ∼100 µL per spray) into both nostrils of nasal spray formulation which contained either 0.1 mg/mL or 0.2 mg/mL of the colostrum immunoglobulin preparation. One hour and four hours after administration, a sample (from either nostril, respectively), was taken using a filter paper piece with a volume capacity of 15 µl which was kept on the nasal mucosa (medial surface of the inferior turbinate) for 10 minutes. As a baseline, a sample from each individual was taken prior to the administration as well. The samples were dissolved in 235 µL PBS buffer containing 0.05% Tween20 and protease inhibitor cocktail (Roche), and analyzed using a Cow IgG ELISA kit (Abcam ab190517). The OD450 values of the baseline samples were subtracted from the OD450 values of the samples collected after nasal spray administration, followed by IgG concentration calculations according to manufacturer’s instructions.

### Statistical analysis

Statistical analyses were performed with GraphPad Prism version 9.1.0. The IC_50_ concentrations for ELISA assay and pseudovirus assay were determined using 4-parameter nonlinear regression.

## Data Availability

Data presented in the manuscript will be made available upon contacting the corresponding author.
The use of bovine colostrum as a prophylactic agent against SARS-CoV-2 has been patented (US patent application no 63/160,833) by Mario Plaas, K. Kogermann, E. Zusinaite, T. Tiirats, B. Aasmae, A. Kavak, V. Poikalainen, L. Lepasalu, S. Piiskop, S. Rom, R. Oltjer, K. Kangro, E. Sankovski, J. M. Gerhold, R. Pert, A. Mannik, A. Planken, A. Tover, M. Kurashin, M. Ustav and M. Ustav Jr.

## Acknowledgements and funding

This study was supported by the funding from Enterprise Estonia grant number 2014-2020.4.02.21-0317.

## Author contribution

K. Kangro, M. Kurašin, E. Žusinaite, R. Pert, V. Poikalainen, L. Lepasalu, S. Piiskop, R. Oltjer, K. Kogermann, Mario Plaas, T. Tiirats, B. Aasmäe, Mihkel Plaas, D. Krinka, E. Talpsep, M. Kadaja, J. M. Gerhold, A. Planken, A. Tover, A. Merits, A. Männik, M. Ustav Jr, M. Ustav designed research; K. Gildemann, E. Sankovski, E. Žusinaite, L. S. Lello, A. Kavak, V. Poikalainen, L. Lepasalu, M. Kuusk, R. Pau, S. Rom, K. Tiirik performed research; K. Kangro, M. Kurašin, E. Žusinaite, E. Sankovski, J. M. Gerhold, A. Planken, A. Männik, M. Ustav Jr analyzed data; K. Kangro, J. M. Gerhold, A. Männik and M. Ustav Jr wrote the paper.

